# Plasma p-tau markers and vascular factors are associated with cognitive decline and clinical progression in the CIMA-Q cohort

**DOI:** 10.1101/2025.09.26.25336739

**Authors:** Rosalie Cottez, Clémence Peyrot, Hélèna L. Denis, Cyntia Tremblay, Andréanne Loiselle, Ali Filali-Mouhim, Sébastien S. Hébert, Nicole Leclerc, Kaj Blennow, Henrik Zetterberg, Consortium for the Early Identification of Alzheimer’s Disease – Quebec (CIMA-Q), Marie-Jeanne Kergoat, Frédéric Calon, Alexa Pichet Binette

## Abstract

**Introduction:** We compared associations between phosphorylated tau biomarkers (p-tau217, p-tau181, p-tau231) and vascular risk factors with clinical progression and cognitive decline along the AD continuum.

**Methods:** Baseline plasma p-tau concentrations and vascular risk factors were assessed in 280 CIMA-Q participants. Associations between these markers, cognition and clinical progression over 8 years were examined.

**Results:** Elevated p-tau217 predicted progression from mild cognitive impairment (MCI) to AD dementia (*p*<.01), while hypertension predicted progression from cognitively unimpaired (CU) to MCI (*p*<.02). Higher p-tau levels, particularly p-tau217, and hypertension were linked to cognitive decline in MCI individuals. In the CU group, few associations were seen between p-tau levels and cognitive decline, with minimal effect of vascular risk factors.

**Discussion:** Plasma p-tau217 was the most sensitive marker of AD-related decline, but hypertension was particularly relevant at earlier stages. The results highlight the need for multimodal profiling to optimize prediction and intervention along the AD continuum.

## 1. BACKGROUND

Alzheimer’s disease (AD) is a multifactorial disease in which pathological changes occur over a long period of time until clinical symptoms appear [1,2]. The core neuropathological markers that define AD are beta-amyloid (Aβ) plaques and tau neurofibrillary tangles [3]. AD pathology can be assessed through positron emission tomography (PET) imaging and cerebrospinal fluid (CSF) analysis [4,5], but these methods remain costly and are not widely accessible. Since a few years, given the fast development of blood-based markers, it is now possible to measure AD pathological biomarkers with high accuracy in the plasma [6]. Prominent markers include phosphorylated tau (p-tau) at different epitopes, such as p-tau181, p-tau231, and most recently p-tau217 [7–12]. Among all markers, p-tau217 has been shown to strongly correlate with AD pathology in the brain, and that has shown the highest diagnostic accuracy for AD in different head-to-head studies [11,13–15]. P-tau231 is thought to be most informative in earlier stages of the disease, with levels increasing before Aβ-PET positivity [16].

Along with AD pathology, clinical trajectories are also influenced by multiple factors [17], among which vascular risk factors (as hypertension and/or high LDL cholesterol) are well established contributors to cognitive decline and increased risk of dementia [18–21]. Together, midlife vascular risk factors might account for up to 10% of AD risk, and multidomain intervention that include improving cardiovascular health is a promising approach to reduce AD risk [22,23]. These comorbidities may interact with AD pathological processes or have an additive effect, ultimately influencing clinical outcomes and highlighting the importance of considering them when evaluating disease progression.

The objective in this study was to evaluate the predictive value of plasma p-tau markers and vascular risk factors on both clinical progression and cognitive decline along the AD continuum over up to 8 years of follow-up data. We hypothesized that the importance of different biological factors might differ depending on whether people are still cognitively unimpaired (CU) versus when they have mild cognitive impairment (MCI), and we thus analyzed the two populations separately. While studies have emerged suggesting the ability of p-tau markers, in particular p-tau217, to track cognitive decline [11,13–15,24], most prior studies had shorter longitudinal data and have not assessed the combination of plasma AD markers and vascular risk factors. To fill this gap, we used data from the Consortium for the Early Identification of Alzheimer’s Disease - Quebec (CIMA-Q), a longitudinal, ongoing cohort of older adults based in the province of Quebec, Canada covering the full continuum of AD [25].

## 2. MATERIALS AND METHODS

### 2.1 Participants

CIMA-Q participants were recruited across the province of Québec between 2019 and 2024 [25,26]. This ongoing consortium aims to facilitate early diagnosis of AD and to identify new therapeutic targets, by ensuring that each participant receives a complete clinical evaluation by a certified physician. Briefly, to be enrolled, participants needed to be at least 65 years old, to understand, read and write French or English, to have an informant available to provide additional information and a Clinical Dementia Rating (CDR, [27,28]) of maximum 1. Exclusion criteria included active substance use disorder, presence or history of a central nervous disorder and regular use of benzodiazepines. Participants were followed up every two years and each visit included a neuropsychological evaluation, a blood draw, questionnaires on psychoaffective measures and comorbidities. At each visit, participants were clinically assessed by expert physicians and underwent a neuropsychological and neuropsychiatric examination with a certified psychometrician. Cognitively unimpaired (CU) participants were recruited from the community. They all had a CDR of 0 and a Montreal Cognitive Assessment (MoCA, [29]) between 26 and 30. They were further characterized as having Subjective Cognitive Decline (SCD) if they were worried that their memory was getting worse [30,31], which formed the majority of the CU group. Patients with AD or some with MCI, were recruited from memory clinics. Participants were diagnosed as MCI according to education-adjusted delayed recall scores on the Wechsler Logical Memory test, a MoCA between 20 and 25, and a CDR score of 0.5. All MCI participants also had to meet NIA/AA clinical criteria for MCI due to AD [31]. Diagnostic criteria for patients with early AD dementia were to have a MoCA between 13 and 24, a CDR score of 1.0, and fulfill NIA/AA clinical criteria for probable AD [31,32]. By design, patients with AD dementia only underwent a baseline visit and were not followed over time.

In cross-sectional analyses, we included all participants who had plasma p-tau measurements available at baseline (n= 283) and longitudinal analyses were restricted to those who had at least one follow-up visit (n= 176). All data was acquired between November 2014 and September 2024. Ethics approval was obtained from the ethics committee of the “Institut Universitaire de Gériatrie de Montréal”. All participants provided a written informed consent in accordance with the Declaration of Helsinki.

### 2.2 Cognitive measures

Participants underwent an extensive neuropsychological evaluation at baseline and at every follow-up visit [25]. Here we focused on 1) the MoCA score as a measure of global cognition (score out of 30); 2) the sum of words remembered from a list over five trials on the Rey Auditory Verbal Learning Test (RAVLT) as a measure of immediate memory (out of 75 as a total score); 3) the number of words remembered from the RAVLT list after a delay as a measure of delayed recall (out of 15); and the score on the Digit Symbol Substitution test (DSST) as a measure of executive function [33].

### 2.3 Vascular risk factors measure

During the medical history assessment, participants reported whether they have ever been diagnosed with hypertension, dyslipidemia or diabetes using a binary format (Yes/No). These comorbidities were tracked at follow-up visits to capture any changes over the last two years. In the current project, only baseline data on these vascular factors were considered.

### 2.4 Plasma p-tau biomarkers

Plasma samples collected at baseline were analyzed for p-tau217, p-tau181, and p-tau231. Plasma p-tau217 levels were measured using the S-PLEX® Human Tau (pT217) kit (MSD, Rockville, MD). Plasma p-tau181 and p-tau231 were assessed using an in-house Single molecule array (Simoa) method at the Clinical Neurochemistry Laboratory, University of Gothenburg, Mölndal, Sweden [8,10]). A total of 283 participants had baseline measurements available for p-tau217, p-tau181 and p-tau231. We excluded three participants who had extreme values on p-tau biomarkers (2 AD and 1 CU), with values exceeding six standard deviations from the mean, resulting in 280 participants retained for analyses. All three later participants had very high creatinine levels, with two of them having the highest level (above 190 µmol/L), which could have also influenced the p-tau measurements [34,35].

### 2.5 CSF biomarkers

A small subset of participants (n = 58) had CSF available at baseline, in which Aβ42, Aβ40 and p-tau181 concentrations were measured. CSF was collected by a neurologist following standard procedures and all lumbar punctures were done in the afternoon. Samples were aliquoted and stored at −80 °C in the CIMA-Q biobank at CHUL (Quebec, QC). Enzyme-linked immunosorbent assays (ELISA) following the manufacturer’s instructions were used to measure Aβ42 and Aβ40 (A Peptide Panel 1 Kit (6E10), #K15200E) and p-tau181 (S-PLEX Human Tau (pT181) Kit, #K151AGMS).

### 2.6 Statistical analysis

Differences in continuous variables between diagnostic groups at baseline were assessed using the Kruskal-Wallis test, followed by Dunn’s post-hoc test. Categorical variables (sex, *APOE* ε4 genotype, hypertension, dyslipidemia and diabetes) were analyzed using the Fisher test. Bivariate associations evaluated how plasma p-tau levels correlated with the CSF measures. First, p-tau levels were compared between diagnostic groups in linear models including age, sex and education as covariates. For each comparison, estimated means adjusted for covariates (marginal means) were extracted. Pairwise group comparisons were performed, and the resulting *P* values were corrected for multiple testing using Tukey’s test.

We next evaluated how plasma p-tau measures at baseline were related to clinical progression over on average 3.10 years of follow-up (range 2 to 8 years), focusing on progression to MCI, and progression to AD dementia in separate Cox proportional hazards models. Time from baseline at which a change in diagnosis occurred was recorded for individuals with clinical progression, and total follow-up time was used for individuals who remained CU or those who remained MCI. A first set of analysis was done including separately each p-tau marker, age, sex and education, followed by another set of analysis further including the vascular risk factors as covariates (hypertension, dyslipidemia, diabetes).

We also investigated associations between p-tau levels and cross-sectional and longitudinal cognitive performance. For cross-sectional associations, linear models were used with baseline measures of p-tau and cognition. For longitudinal analyses, linear mixed-effect models with random slope and intercept were used to assess the effect of baseline p-tau levels and cognitive decline over time. The time variable was the years from baseline. Similar to the analyses related to clinical progression, a first set of models focused on each p-tau marker, including age, sex and education as covariates, and a second set of models further included the vascular risk factors. The analyses were conducted on the whole group, as well as separately in CU and cognitively impaired (CI) individuals, the latter including patients with MCI and AD dementia. Statistical significance for all models was set at p < .05 All analyses were performed in R v.4.4.1 (R Core Team, 2024).

## 3. RESULTS

### 3.1 Sample demographics and associations with plasma p-tau markers

We included 280 participants from the CIMA-Q cohort with available plasma p-tau measurements at baseline, consisting of 168 CU older adults (112 SCD; 56 cognitively normal), 84 patients with MCI and 28 patients with AD dementia. On average, participants were 74.3 years old, had 15.0 years of education, 64% were women, and close to half of the cohort presented vascular risk factors. Demographic, diagnosis and biomarkers information according to clinical diagnosis are shown in **Table 1**.

**Table 1.** Description of the CIMA-Q cohort.

| Characteristics | CU<br>(n=168) | MCI<br>(n=84) | AD<br>(n=28) | ALL<br>(n=280) | P value |
| --- | --- | --- | --- | --- | --- |
| **Clinical and sociodemographic data** |  |  |  |  |  |
| Age (years) | 72.8 (5.4) | 76.3 (6.1) | 76.8 (6.5) | 74.3 (6.0) | **<.001** |
| Gender (Women, %) | 123 (73 %) | 41 (49 %) | 14 (50 %) | 178 (64 %) | **<.001** |
| BMI (kg/m<sup>2</sup>) | 26.5 (4.6) | 27.1 (4.4) | 26.1 (3.5) | 26.6 (4.4) | .27 |
| *APOE* ε4 carriers<br>(≥1 allele, %) | 37 (22 %) | 30 (36 %) | 14 (50 %) | 81 (29 %) | **.003** |
| Hypertension, n(%)<sup>a</sup> | 62 (39 %) | 42 (54 %) | 10 (42 %) | 114 (43 %) | .08 |
| Dyslipidemia, n(%)<sup>b</sup> | 63 (39 %) | 44 (56 %) | 11 (48 %) | 118 (45 %) | **.04** |
| Diabetes, n(%) | 14 (8.3 %) | 11 (13 %) | 1 (3.6 %) | 26 (9.3 %) | .31 |
| Education (years)<sup>c</sup> | 15.1 (3.6) | 14.6 (3.7) | 15.3 (4.8) | 15.0 (3.8) | .44 |
| Follow-up time (years) | 3.3 (1.0) | 2.9 (1.1) | \ | 3.1 (1.0) | **<.001** |
| **Cognition** |  |  |  |  |  |
| MoCA | 27.2 (2.2) | 23.7 (2.6) | 19.1 (4.2) | 25.3 (3.7) | **<.001** |
| RAVLT total score | 46.8 (9.1) | 35.4 (10.4) | 22.9 (5.8) | 41.0 (12.1) | **<.001** |
| RAVLT delayed recall<sup>d</sup> | 9.8 (2.9) | 5.8 (3.7) | 1.4 (1.9) | 7.8 (4.1) | **<.001** |
| Digit symbol | 60.4 (12.2) | 48.9 (15.7) | 40.3 (13.7) | 55.1 (15.2) | **<.001** |
| **Plasma p-tau biomarkers** |  |  |  |  |  |
| p-tau217 (pg/mL) | 3.2 (2.5) | 4.7 (3.3) | 10.8 (6.0) | 4.4 (3.9) | **<.001** |
| p-tau181 (pg/mL) | 5.0 (2.6) | 6.6 (3.2) | 10.3 (5.0) | 6.0 (3.5) | **<.001** |
| p-tau231 (pg/mL) | 5.9 (2.3) | 7.0 (2.8) | 9.4 (3.5) | 6.6 (2.8) | **<.001** |

*Note:* All values presented are means (±SD) unless otherwise specified. AD participants were not followed over time (\). *AD*, Alzheimer's disease; *APOE*, Apolipoprotein E; *BMI*, body mass index; *CU*, cognitively unimpaired; *MCI*, mild cognitive impairment; *MoCA*, Montreal Cognitive Assessment; *RAVLT*, Rey Auditory Verbal Learning Test. Missing data: <sup>a</sup> = 17, <sup>b</sup> = 18, <sup>c</sup> = 1, <sup>d</sup> = 6, <sup>e</sup> = 4

Regarding plasma biomarkers, p-tau217, p-tau181, and p-tau231 levels were higher in AD patients compared to MCI or CU participants (all p < .0001), particularly for p-tau217 (marginal mean AD-CU = 1.80 (SE .17) and AD-MCI = 1.53 (.18)) compared to p-tau181 (AD-CU = 1.33 (.18) and AD-MCI = 1.07 (.19)) or p-tau231 (AD-CU = 1.12 (.19) and AD-MCI = 0.86 (.20)).

Also, there was a trend toward higher p-tau217 in MCI compared to CU participants (MCI-CU = 0.27 (.12); p = .06) (**Figure 1A**). Dyslipidemia was the main vascular factor that differed between diagnostic groups (*p* = .04, **Table 1**) and was most prevalent in participants with MCI (56%).

**Figure 1.**
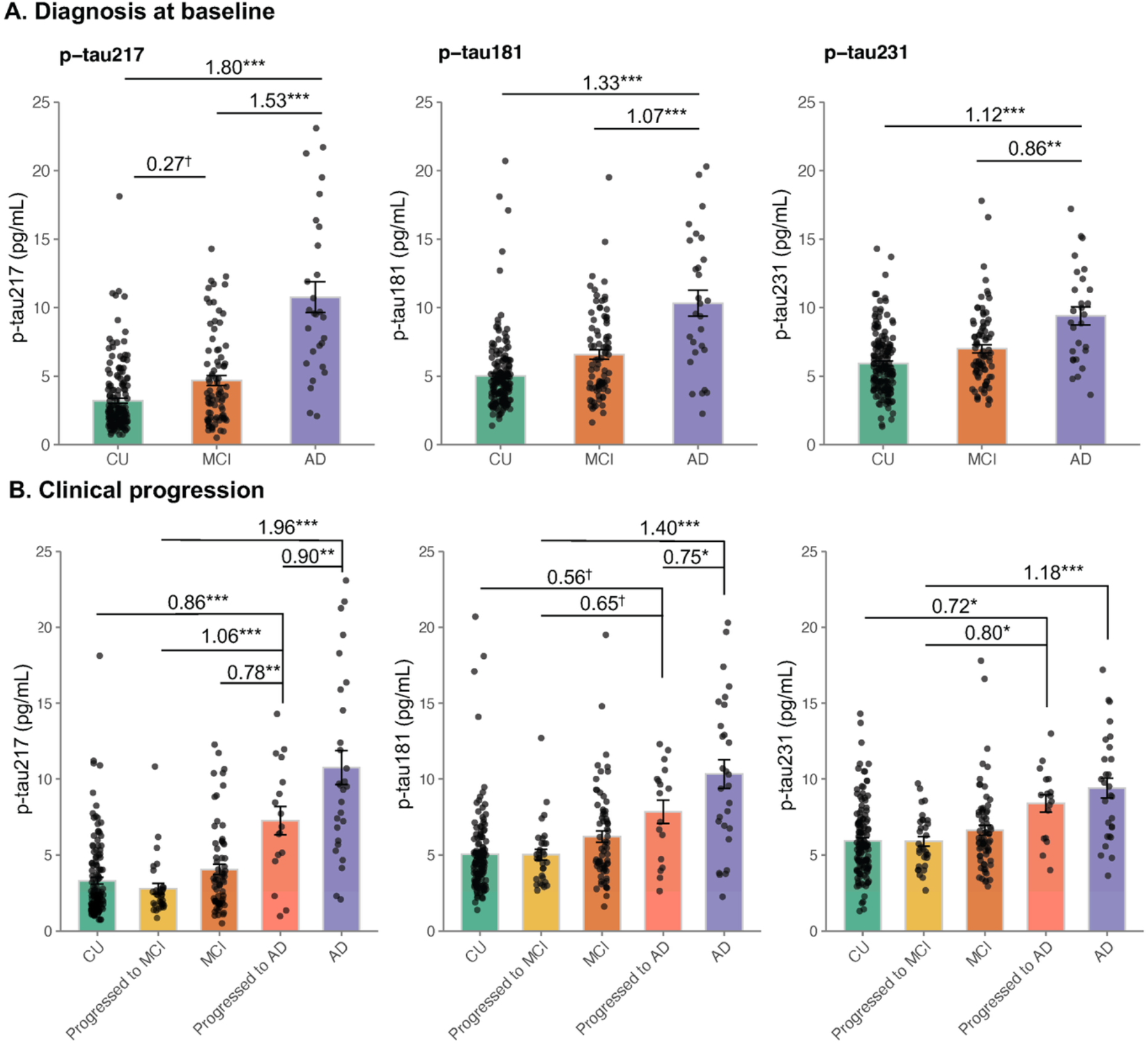
Plasma p-tau217, p-tau181 and p-tau231 levels by clinical diagnosis. **A.** Plasma p-tau levels by clinical diagnosis at baseline. **B**. Plasma p-tau levels incorporating clinical progression. To simplify the visualization, only comparisons involving at least one progressor group are shown (**B**). In all panels, standardized marginal means comparing diagnostic groups, adjusted for age, sex and education, are reported. *P* values were adjusted for multiple comparisons with the Tukey’s test. Bars indicate mean (±SE). *AD, Alzheimer’s disease; CU; Cognitively unimpaired; Progressed to MCI, CU participants at baseline who progressed to MCI; MCI, mild cognitive impairment; Progressed to AD, MCI participants at baseline who progressed to AD dementia; p-tau, phosphorylated tau*. Statistical significance is indicated as follows: ^†^ trend-level *p* < .10, * *p* < .05; ** *p* < .01; *** *p* < .001.

In the subset of participants with CSF measurements (n = 58), all plasma p-tau measurements correlated with CSF p-tau181, with the highest correlations seen with plasma p-tau217 (0.84 compared to 0.48 and 0.49 with p-tau181 and p-tau231, all *p* < .001). Lower correlations were seen with CSF Aβ42/40 for all plasma p-tau markers (−0.53 to −0.37) (**Supplementary Figure 1**).

### 3.2 Associations between p-tau markers and clinical progression

Next, we investigated how the baseline plasma p-tau markers related to subsequent clinical progression over an average period of 3.10 years. Looking at clinical progression to MCI, there were no differences in the three p-tau markers between the CU participants who progressed to MCI (n = 32 out of 112) and those who remained CU (**Figure 1B**, all *p* > .71). Looking at clinical progression to AD, the patients with MCI who progressed to AD dementia (n = 17 out of 64) had higher concentrations of p-tau217 at baseline compared to those who remained with a diagnosis of MCI (**Figure 1B**, marginal mean progAD-MCI = 0.78 (SE .22), *p* = .003), and no differences between groups were seen for p-tau181 or p-tau231 (progAD-MCI = 0.41 (.23) and 0.59 (.25), *p* = .40 and .12, respectively).

Such results were recapitulated in survival analyses using Cox proportional hazards model (**Figure 2**). In the CU group, none of the p-tau markers were associated with progression to MCI (hazard ratio [HR] between 0.97 and 1.04, all *p* > .53). In the MCI group, higher p-tau217 levels at baseline was associated with progression to AD (HR = 1.22, *p* = .007 for continuous p-tau levels). In splitting p-tau217 levels into tertiles for visualization and to ease interpretation of risk of progression, we showed that participants in the top tertile had higher risk of progressing to AD dementia compared to the two other groups: HR = 6.67 (*p* = .02) and 4.25 (*p* = .04) between the group of participants in the highest tertile of p-tau217 concentrations compared to the first and second tertile, respectively (**Figure 2A-B**). No differences between tertiles on any of the p-tau markers were seen in relation to progression to MCI.

**Figure 2.**
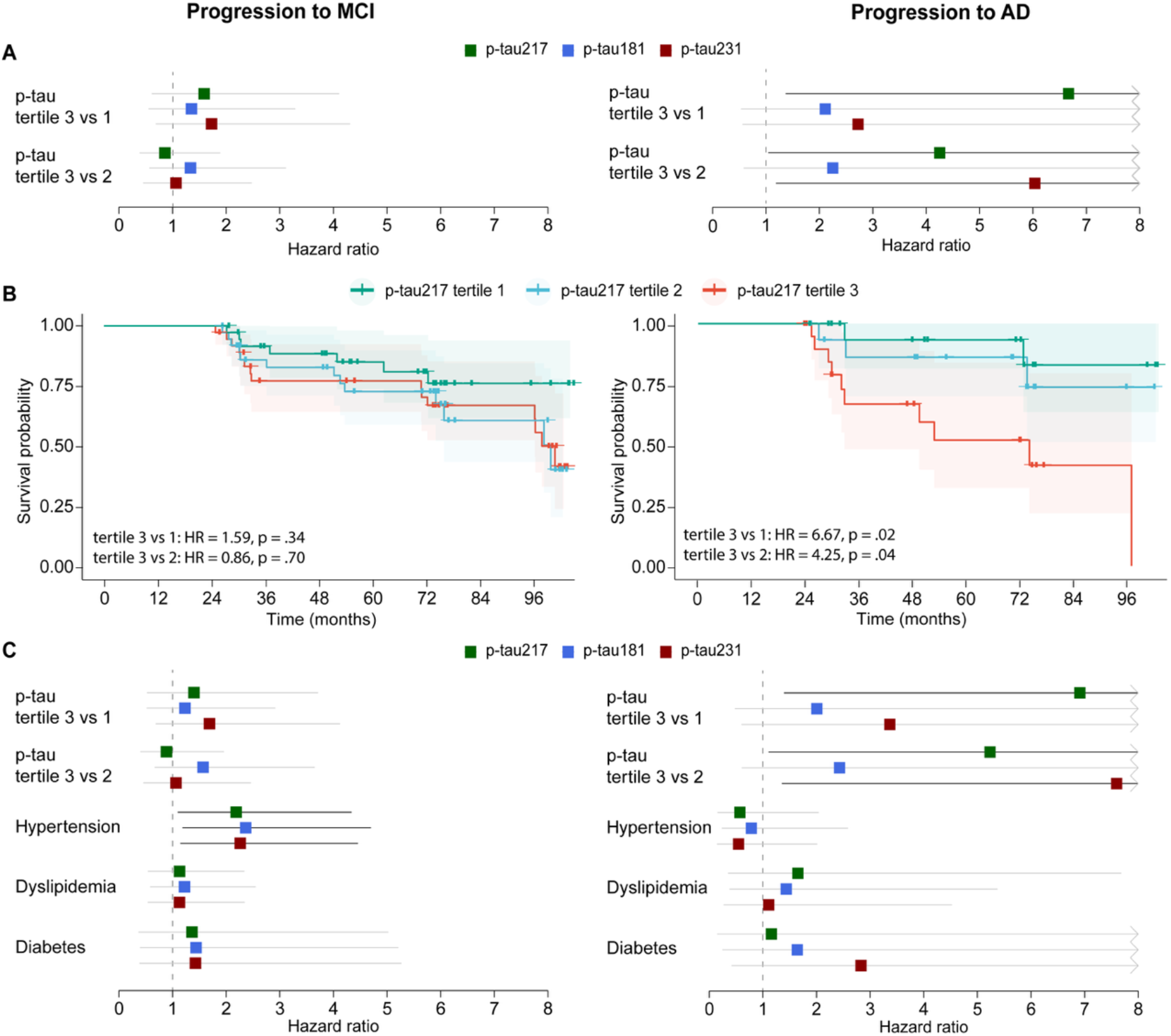
Clinical progression to MCI or AD. **A.** Forest plots showing the hazard ratios (HR) and 95% confidence intervals (CIs) derived from Cox regression models using the highest tertile (tertile 3) of p-tau concentration as the reference. Separate models were fitted with each p-tau marker, including age, sex and education as covariates. **B**. Survival curves illustrating the progression to MCI or AD based on tertiles of p-tau217. Comparisons are made between tertiles 1 and 2 to tertile 3, separately for CU participants who progressed to MCI (left) and for participants with MCI who progressed to AD dementia (right). **C**. Forest plots showing the hazard ratios (HR) and 95% confidence intervals (CIs) derived from Cox regression models when further including vascular risk factors. Separate models were fitted with each p-tau marker, along with hypertension, dyslipidemia, diabetes, age, sex and education. The dashed line indicates a HR of 1. The darker CI lines indicate significant associations. *AD, Alzheimer’s disease; MCI, Mild cognitive impairment; p-tau, phosphorylated tau*.

### 3.3 Additional markers associated with clinical progression

Given the relatively low levels of baseline plasma p-tau markers in individuals who progressed to MCI, we further investigated what might be other measures influencing clinical progression. Our analysis focused on vascular risk factors, i.e. self-reported hypertension, dyslipidemia and diabetes, as important risk factors of age-related cognitive impairment. In the CU group, including all factors in a common model with each p-tau measure, hypertension was the main factor associated with progression to MCI (HR between 2.29 and 2.32, all *p* < .02; **Figure 2C**).

None of the other factors were significant. In the MCI group, when including the different vascular risk factors, higher plasma p-tau217 levels at baseline remained the only measure associated with progression to AD (HR = 1.25, *p* = .006 for continuous levels and tertile measures are reported on **Figure 2C**). All statistical details are reported in **Supplementary Table 1** for progression to MCI and **Supplementary Table 2** for progression to AD dementia.

### 3.4 Associations between p-tau markers and cognition

We next investigated associations between plasma p-tau markers, vascular risk factors and cognitive performance at baseline and over time (**Figure 3**). In cross-sectional analyses among CU individuals, none of the p-tau markers were associated with cognitive performance. When vascular risk factors were added to the models, hypertension emerged as the only factor significantly associated with lower score on delayed recall (std β = −0.20 and −0.19, all *p* < .02; **Figure 3A**). In contrast, among CI participants, cross-sectional analyses revealed consistent negative associations between all p-tau markers and cognitive test scores. Notably, the strongest association was between p-tau217 and the MoCA (std β = −0.37 for p-tau217 and −0.22 for the other p-tau’s; **Figure 3B**). Additional adjustment for vascular risk factors led to attenuation of several associations, particularly those related to digit-symbol substitution (**Figure 3A**).

**Figure 3.**
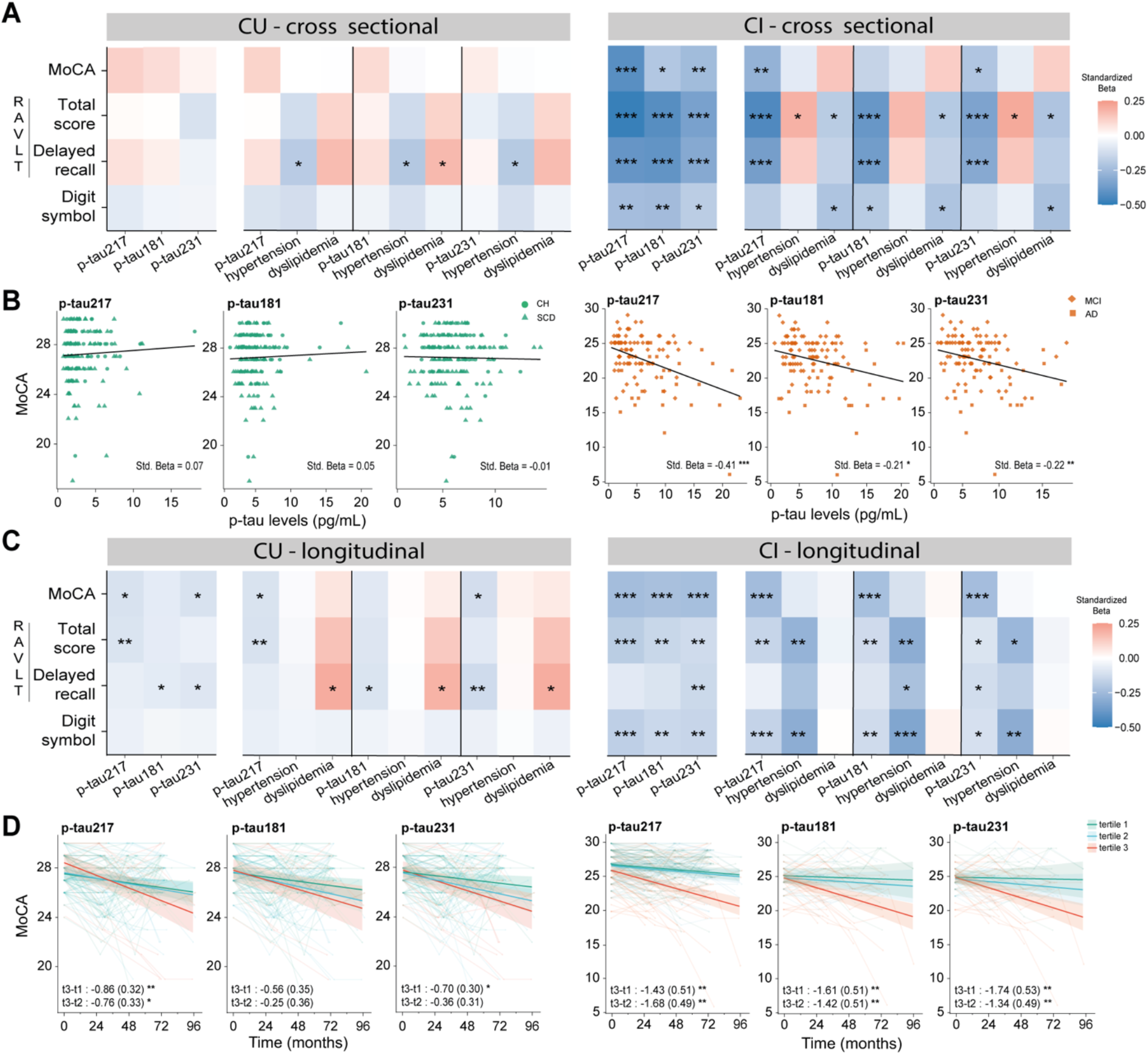
Associations at baseline and over eight years between p-tau markers and cognition. **A.** Heatmaps showing associations between plasma p-tau levels and vascular risk factors, with cognitive performance in Cognitively Unimpaired (left) and Cognitively Impaired (right) participants for cross-sectional associations. **B**. Scatter plots showing associations between individual plasma p-tau levels and MoCA performance at baseline in Cognitively unimpaired (left) and Cognitively Impaired (right). **C**. Heatmaps showing associations between plasma p-tau levels and vascular risk factors at baseline on cognitive decline. Estimates reported correspond to the interaction between time and the marker of interest. All p-tau measures were included as continuous values **D**. For visualization purposes, participants were grouped into tertiles to show the effect of baseline p-tau levels on MoCA performance over time. *AD, Alzheimer’s disease; CI, cognitive impairment; CU; Cognitively unimpaired; MCI, mild cognitive impairment; MoCA, Montreal Cognitive Assessment;* p-tau, phosphorylated tau; *RAVLT, Rey Auditory Verbal Learning Test; t, tertile (1, 2 or 3)*. Statistical significance is indicated as follows: * *p* < .05; ** *p* < .01; *** *p* < .001.

In longitudinal analyses, higher levels of p-tau217 and p-tau231 at baseline were significantly associated with greater decline on the MoCA over time in CU individuals (both estimates = −0.10, *p* < .05). A few additional associations were observed between p-tau markers and decline on memory performance on the RAVLT (**Figure 3C**). These associations remained robust after adjusting for vascular risk factors. All statistical associations are reported in **Supplementary Table 3**. Among CI participants, higher baseline levels of all p-tau markers were associated with longitudinal decline across most cognitive domains. The highest effect was again observed with the MoCA, with a more pronounced association for p-tau217 (p-tau217*time = −0.24, *p* < .01). These patterns remained unchanged following adjustment for vascular risk factors. While p-tau markers remained the primary predictors of cognitive decline, hypertension was also independently associated with steeper decline on approximately half of the cognitive measures in the CI group. Specifically, significant hypertension*time interactions were observed for RAVLT total score and digit-symbol substitution, with estimates ranging from −0.27 to −0.33 (all *P* values < .01, **Figure 3C**). All statistical associations are reported in **Supplementary Table 4**. To illustrate the longitudinal effects on the MoCA, participants were stratified by tertiles of p-tau levels (**Figure 3D**). Overall, individuals in the highest tertiles of p-tau217 and also p-tau231, in both CU and CI subgroups declined over time.

Similar patterns as those seen in CI were observed in the whole cohort (**Supplementary Figure 2)**. As expected, higher baseline levels of all plasma p-tau markers were consistently associated with lower cognitive performance at baseline and greater cognitive decline over time across all cognitive measures. Vascular risk factors mainly influenced cognitive decline, with hypertension emerging as a significant predictor of worsening cognitive outcomes over time (**Supplementary Figure 2)**.

## DISCUSSION

In this longitudinal investigation across the AD continuum in the CIMA-Q cohort, we integrated plasma p-tau measures and vascular risk factors to better understand cognitive decline and clinical progression over 8 years. Elevated plasma p-tau217 levels were associated with progression from MCI to AD dementia and with accelerated cognitive decline, particularly in CI individuals. Hypertension emerged as the only significant vascular risk factor. In the CU group, who on average had relatively low p-tau levels, hypertension was the only marker of progression to MCI. By contrast, in the CI group, both p-tau levels and hypertension contributed to cognitive decline, suggesting that the relative importance of these markers may shift depending on cognitive stage and underlying levels of AD pathology. These findings highlight the value of considering both vascular risk factors and AD biomarkers for risk stratification and tailored prevention or intervention strategies to reduce dementia risk.

Plasma p-tau217 levels, and to a lesser extent p-tau181, were significantly associated with faster cognitive decline across the entire cohort, and more strongly in CI individuals, in line with recent findings comparing plasma p-tau217, p-tau181 and p-tau231 [36]. In CU participants, none of the p-tau markers were significantly associated with cognitive scores and clinical progression, which differed from some previous studies [37–39]. Most of the CU participants who progressed to MCI had low levels of p-tau217 (most below 3.6 pg/mL, corresponding to the middle tertile, **Figure 1B**) and fell below the recently proposed Aβ-positivity cut-offs between 5.0-6.0 pg/mL using the same assay [40]. This suggests that overall there is a low proportion of individuals in the preclinical phase of AD and (an)other etiology(ies) than AD likely underly the clinical progression, highlighting the importance of examining other risk factors. Still, even though the CU group exhibited modest cognitive decline, individuals with the highest p-tau217 and p-tau231 levels showed faster cognitive decline over time, highlighting the predictive value of those plasma markers before the appearance of symptoms.

These individuals might still be years away from clinical progression, and longer follow-up will help clarify their trajectories.

Among the other biological factors, hypertension emerged as the only vascular factor associated with progression to MCI, consistent with prior evidence showing that mid- and late-life hypertension increases risk of cognitive decline, whereas blood pressure reduction mitigates it, as shown in a large population-based cohort study [41] and clinical trial [42]. These results reinforce the idea that hypertension is an important risk factor that influences progression to MCI in older adults. In relation to cognitive decline, in the whole group as well as in CI participants, both p-tau levels and hypertension were both associated with worse performance over time, further confirming hypertension as an important risk factor of cognitive impairment. Links between hypertension and AD pathology had been proposed recently, with higher blood pressure being related to higher Aβ PET burden and accumulation over time [43]. We speculate that the presence of hypertension and AD pathology might have an additive effect on clinical progression and cognitive decline [20,44] as well as independent effect. As noted above, in the CU group that presented low p-tau levels, hypertension emerged as the only factor explaining frank progression to MCI, suggesting it is an important independent factor leading to MCI likely not due to AD. Overall, in this context, our results emphasize the importance of integrating established AD biomarkers with measures of blood pressure to refine early risk prediction and increase opportunities to slow disease progression, especially since hypertension can be treated efficiently [42,50,51].

Our study has both strengths and limitations. The long follow-up period (up to 8 years) allowed us to identify and track individuals who progressed along the AD continuum, providing valuable longitudinal insight. While we cannot confirm the presence of AD pathology using PET or other methods, all clinical diagnoses are robust and determined by a physician after a comprehensive clinical evaluation. The CIMA-Q cohort is also not enriched for individuals at higher risk of AD (e.g., family history, *APOE*4 genotype screening5.0). Despite the modest sample size (n = 280), our results complement effects observed in larger studies within the field and highlight the importance of combining multiple markers to better understand cognitive decline and clinical progression. Based on the p-tau217 levels, the overall proportion of CU individuals who are likely to be Aβ-positive seems lower than in some previous large-scale studies focusing on the asymptomatic phase of the disease [24,38,52], which likely explains some of the lower associations with cognition and clinical progression with p-tau217 than previously reported. The cohort has limited ethnic diversity – participants are predominantly White French-Canadians– which may also restrict the applicability of findings to more heterogeneous populations. This is important as plasma p-tau levels [53,54] and vascular burden [55,56] might differ based on ethnic groups.

In conclusion, by integrating plasma p-tau biomarkers and vascular risk factors in relation to both clinical progression and cognitive trajectories, our study contributes to refining p-tau217 and hypertension as accessible and important measures for AD progression. Plasma p-tau217 is a valuable addition to individualized monitoring frameworks, particularly when interpreted alongside vascular and clinical risk profiles.

## Data Availability

All data produced in the present study are available upon reasonable request to the authors

## ACKNOWLEDGEMENTS

The data used in this article were obtained from the Consortium pour l’identification précoce de la maladie Alzheimer - Québec (CIMA-Q; cima-q.ca). The investigators within the CIMA-Q contributed to the design, the implementation of the research, the recruitment and follow-up, the acquisition of clinical, cognitive and biological samples. A list of the CIMA-Q investigators is available on www.cima-q.ca

## CONSENT STATEMENT

All participants provided written informed consent.

