## Supplementary material for "Plasma p-tau markers and vascular factors are associated with cognitive decline and clinical progression in the CIMA-Q cohort"

**Supplementary Table 1.** Statistical analyses for progression to MCI

**Supplementary Table 2.** Statistical analyses for progression to AD dementia

**Supplementary Table 3.** Statistical analyses for associations with longitudinal cognition in the CU group

**Supplementary Table 4.** Statistical analyses for associations with longitudinal cognition in the CI group

**Supplementary Figure 1.** Associations between plasma and CSF biomarkers

**Supplementary Figure 2.** Associations with cognition in the whole group

**Supplementary Table 1.** Statistical analyses for progression to MCI

|  | **p-tau217** | | **p-tau181** | | **p-tau231** | |
| --- | --- | --- | --- | --- | --- | --- |
|  | HR (SE) | p | HR (SE) | p | HR (SE) | p |
| p-tau | 0.97 (0.08) | 0.74 | 1.04 (0.07) | 0.53 | 1.05 (0.08) | 0.57 |
| Age | 1.06 (0.03) | 0.05 | 1.06 (0.03) | 0.08 | 1.05 (0.03) | 0.10 |
| Sex (men) | 0.60 (0.39) | 0.19 | 0.66 (0.40) | 0.30 | 0.66 (0.39) | 0.28 |
| Education | 0.94 (0.05) | 0.23 | 0.93 (0.06) | 0.18 | 0.93 (0.06) | 0.19 |
| Model with vascular risk factors | | | | | | |
| p-tau | 1.00 (0.08) | 0.99 | 1.07 (0.07) | 0.37 | 1.06 (0.08) | 0.48 |
| Age | 1.06 (0.03) | 0.07 | 1.05 (0.03) | 0.10 | 1.05 (0.03) | 0.10 |
| Sex (men) | 0.68 (0.40) | 0.33 | 0.73 (0.40) | 0.43 | 0.71 (0.39) | 0.37 |
| Education | 0.94 (0.05) | 0.22 | 0.93 (0.05) | 0.17 | 0.93 (0.05) | 0.18 |
| Hypertension | **2.29 (0.35)** | **0.02** | **2.32 (0.35)** | **0.01** | **2.29 (0.35)** | **0.02** |
| Dyslipidemia | 1.15 (0.37) | 0.71 | 1.19 (0.38) | 0.64 | 1.17 (0.37) | 0.68 |
| Diabetes | 1.38 (0.66) | 0.62 | 1.40 (0.66) | 0.61 | 1.43 (0.66) | 0.59 |

Hazard ratios (HR) and standard error (SE) derived from Cox regression models are shown. Separate models were fitted with each p-tau marker, along with hypertension, dyslipidemia, diabetes, age, sex and education. Significant associations are bolded.

**Supplementary Table 2.** Statistical analyses for progression to AD dementia

|  | **p-tau217** | | **p-tau181** | | **p-tau231** | |
| --- | --- | --- | --- | --- | --- | --- |
|  | HR (SE) | p | HR (SE) | p | HR (SE) | p |
| p-tau | **1.22 (0.07)** | **0.01** | 1.14 (0.08) | 0.11 | 1.09 (0.08) | 0.25 |
| Age | 1.05 (0.05) | 0.32 | 1.07 (0.05) | 0.18 | 1.07 (0.05) | 0.18 |
| Sex (men) | 0.71 (0.59) | 0.55 | 1.25 (0.53) | 0.67 | 1.02 (0.55) | 0.97 |
| Education | 0.88 (0.09) | 0.15 | 0.89 (0.09) | 0.19 | 0.89 (0.09) | 0.16 |
| Model with vascular risk factors | | | | | | |
| p-tau | **1.25 (0.08)** | **0.01** | 1.14 (0.08) | 0.11 | 1.10 (0.08) | 0.26 |
| Age | 1.06 (0.07) | 0.35 | 1.07 (0.06) | 0.26 | 1.07 (0.06) | 0.25 |
| Sex (men) | 0.76 (0.63) | 0.66 | 1.34 (0.57) | 0.61 | 1.11 (0.58) | 0.86 |
| Education | 0.88 (0.09) | 0.15 | 0.89 (0.09) | 0.20 | 0.89 (0.09) | 0.18 |
| Hypertension | 0.64 (0.60) | 0.45 | 0.80 (0.60) | 0.71 | 0.72 (0.63) | 0.60 |
| Dyslipidemia | 1.47 (0.71) | 0.58 | 1.32 (0.71) | 0.69 | 1.34 (0.68) | 0.67 |
| Diabetes | 1.92 (1.00) | 0.51 | 1.79 (0.98) | 0.55 | 1.63 (0.96) | 0.61 |

Hazard ratios (HR) and standard error (SE) derived from Cox regression models are shown. Separate models were fitted with each p-tau marker, along with hypertension, dyslipidemia, diabetes, age, sex and education. Significant associations are bolded.

**Supplementary Table 3.** Statistical analyses for associations with longitudinal cognition in the CU group

|  | p-tau217 | | p-tau181 | | p-tau231 | |
| --- | --- | --- | --- | --- | --- | --- |
|  | Estimate | p | Estimate | p | Estimate | p |
| **MoCA** |  |  |  |  |  |  |
| p-tau | **-0.10** | **0.03** | -0.08 | 0.12 | -0.12 | **0.03** |
| Hypertension | -0.02 | 0.87 | 0.01 | 0.94 | 0.03 | 0.76 |
| Dyslipidemia | 0.07 | 0.52 | 0.04 | 0.68 | 0.05 | 0.61 |
| Diabetes | -0.04 | 0.83 | 0.00 | 0.98 | -0.04 | 0.84 |
| **RAVLT** |  |  |  |  |  |  |
| **Total score** |  |  |  |  |  |  |
| p-tau | **-0.10** | **0.01** | -0.09 | 0.07 | -0.08 | 0.13 |
| Hypertension | -0.03 | 0.77 | 0.00 | 0.96 | 0.02 | 0.86 |
| Dyslipidemia | 0.15 | 0.09 | 0.13 | 0.14 | 0.14 | 0.10 |
| Diabetes | -0.10 | 0.56 | -0.06 | 0.71 | -0.12 | 0.49 |
| **Delayed recall** |  |  |  |  |  |  |
| p-tau | -0.07 | 0.07 | **-0.10** | **0.03** | **-0.13** | **0.01** |
| Hypertension | -0.04 | 0.67 | -0.01 | 0.89 | 0.01 | 0.86 |
| Dyslipidemia | **0.20** | **0.02** | 0.18 | 0.03 | 0.20 | 0.02 |
| Diabetes | -0.08 | 0.62 | -0.02 | 0.88 | -0.08 | 0.63 |
| **Digit symbol** |  |  |  |  |  |  |
| p-tau | -0.06 | 0.12 | -0.04 | 0.33 | -0.05 | 0.27 |
| Hypertension | -0.04 | 0.63 | -0.02 | 0.76 | -0.01 | 0.85 |
| Dyslipidemia | -0.05 | 0.56 | -0.05 | 0.48 | -0.05 | 0.53 |
| Diabetes | -0.03 | 0.85 | -0.02 | 0.91 | -0.04 | 0.81 |

Linear mixed-effect models with random slope and intercept. Estimates reported correspond to the interaction between time and the marker of interest, with age, sex and education also included as covariates in models. All p-tau measures were included as continuous values. *MoCA, Montreal Cognitive Assessment; p-tau, phosphorylated tau; RAVLT, Rey Auditory Verbal Learning Test; SCD, subjective cognitive decline*

**Supplementary Table 4.** Statistical analyses for associations with longitudinal cognition in the CI group

|  | p-tau217 | | p-tau181 | | p-tau231 | |
| --- | --- | --- | --- | --- | --- | --- |
|  | Estimate | p | Estimate | p | Estimate | p |
| **MoCA** |  |  |  |  |  |  |
| p-tau | **-0.24** | **<0.001** | **-0.23** | **<0.001** | **-0.24** | **<0.001** |
| Hypertension | -0.08 | 0.56 | -0.11 | 0.41 | -0.02 | 0.87 |
| Dyslipidemia | -0.04 | 0.79 | 0.02 | 0.91 | 0.00 | 0.97 |
| Diabetes | -0.16 | 0.50 | -0.21 | 0.39 | -0.17 | 0.47 |
| **RAVLT** |  |  |  |  |  |  |
| **Total score** |  |  |  |  |  |  |
| p-tau | **-0.15** | **<0.001** | **-0.13** | **<0.001** | **-0.10** | **0.05** |
| Hypertension | **-0.28** | **<0.001** | **-0.30** | **<0.001** | **-0.26** | **0.01** |
| Dyslipidemia | -0.04 | 0.64 | 0.00 | 0.99 | -0.03 | 0.73 |
| Diabetes | 0.15 | 0.41 | 0.11 | 0.55 | 0.14 | 0.44 |
| **Delayed recall** |  |  |  |  |  |  |
| p-tau | -0.05 | 0.34 | -0.09 | 0.08 | **-0.12** | **0.01** |
| Hypertension | **-0.22** | **0.05** | **-0.22** | **0.04** | -0.17 | 0.10 |
| Dyslipidemia | -0.02 | 0.83 | 0.00 | 1.00 | -0.02 | 0.84 |
| Diabetes | 0.32 | 0.13 | 0.29 | 0.16 | 0.31 | 0.11 |
| **Digit symbol** |  |  |  |  |  |  |
| p-tau | **-0.16** | **<0.001** | **-0.13** | **0.01** | **-0.11** | **0.04** |
| Hypertension | **-0.30** | **<0.001** | **-0.33** | **<0.001** | **-0.30** | **<0.001** |
| Dyslipidemia | 0.01 | 0.95 | 0.03 | 0.74 | 0.01 | 0.92 |
| Diabetes | 0.11 | 0.49 | 0.07 | 0.69 | 0.11 | 0.57 |

Linear mixed-effect models with random slope and intercept. Estimates reported correspond to the interaction between time and the marker of interest, with age, sex and education also included as covariates in models. All p-tau measures were included as continuous values. *MoCA, Montreal Cognitive Assessment; p-tau, phosphorylated tau; RAVLT, Rey Auditory Verbal Learning Test; SCD, subjective cognitive decline*

**Supplementary Figure 1.** Associations between plasma and CSF biomarkers

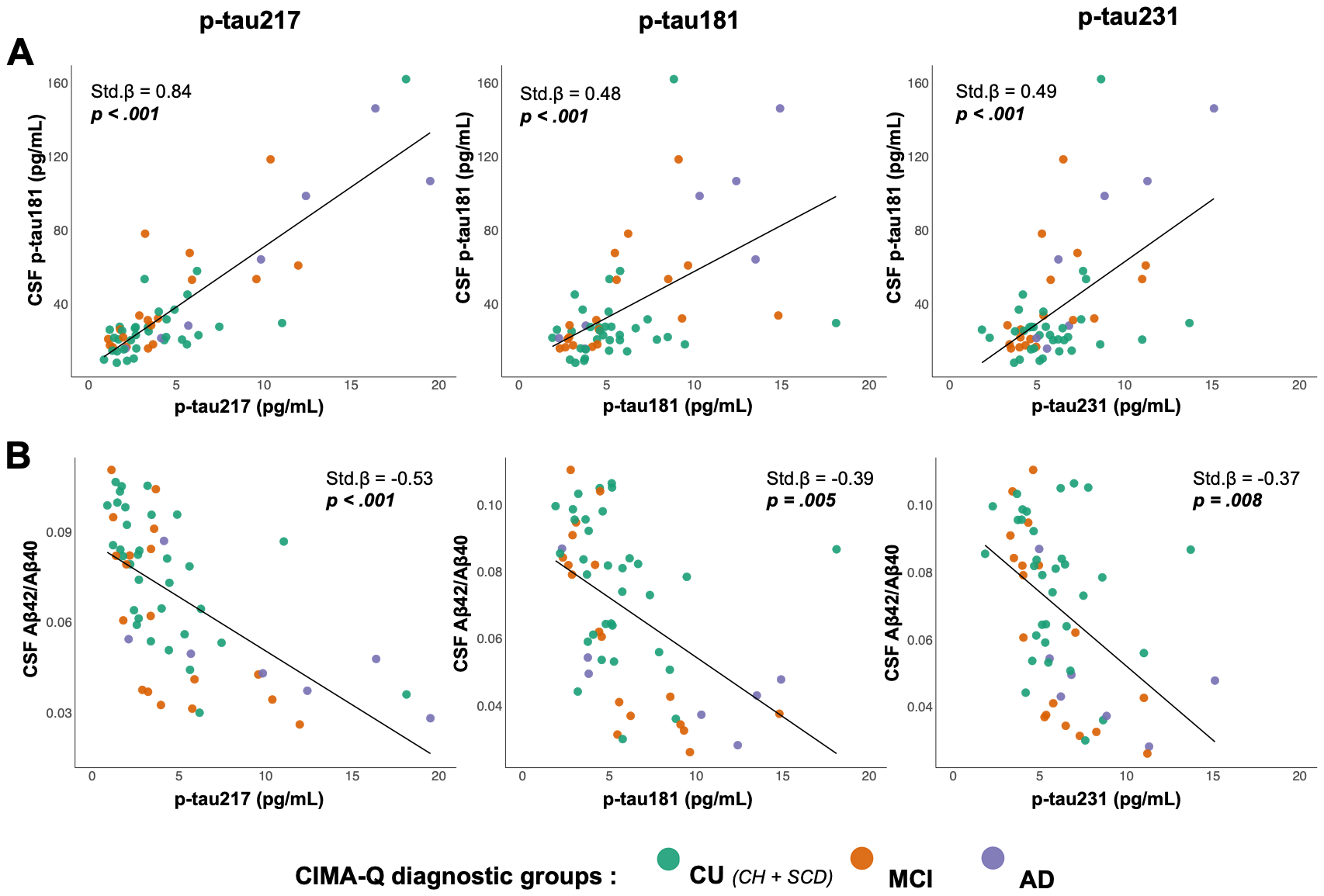

**A**. Associations between plasma p-tau markers and CSF p-tau181. **B**. Associations between plasma p-tau markers and CSF Aβ42/Aβ40. Linear models adjusted for age, sex and education were used with baseline measures of p-tau and cognition. Standardized beta (Std β) coefficients are shown. *Aβ, amyloid-beta peptide; AD, Alzheimer’s disease; CSF, cerebrospinal fluid; CU, cognitively unimpaired; MCI, mild cognitive impairment.*

**Supplementary Figure 2.** Associations with cognition in the whole group

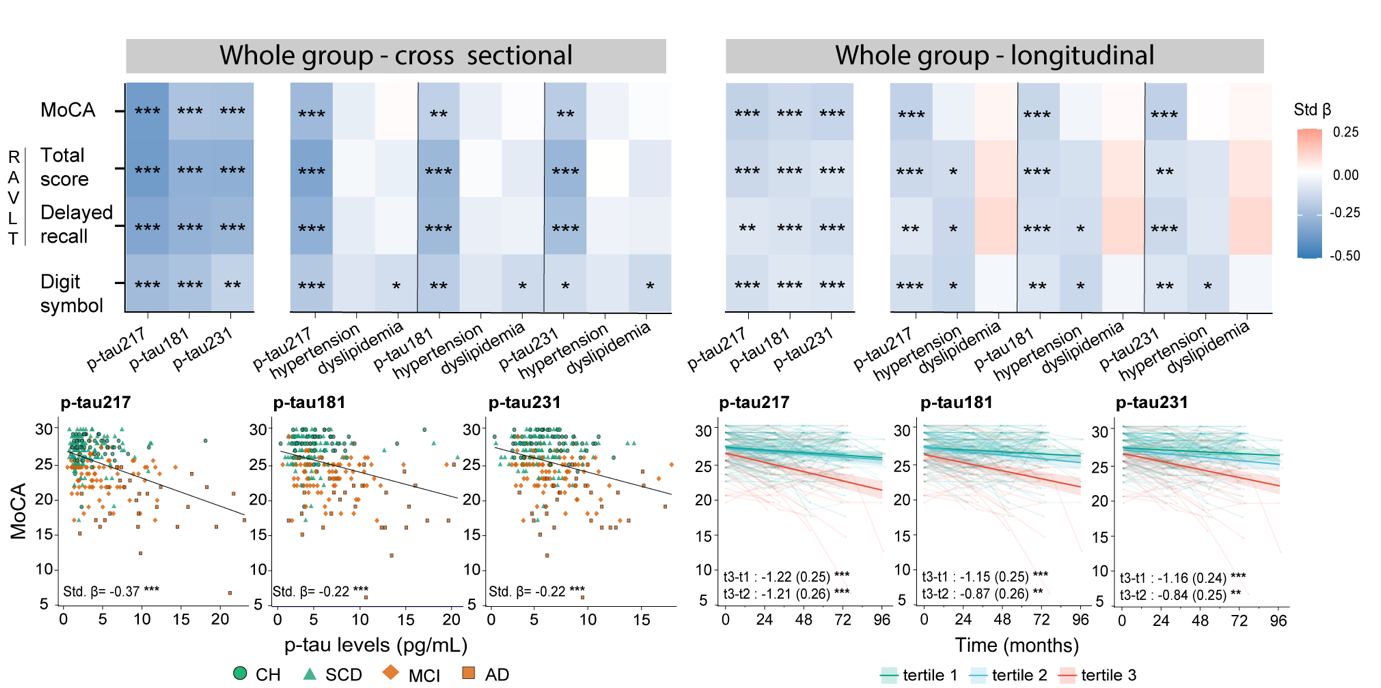

**A.** Heatmap showing associations between plasma p-tau levels and vascular risk factors, with cognitive performance in the whole group for cross-sectional associations. **B.** Scatter plots showing associations between individual plasma p-tau levels and MoCA performance at baseline. **C.** Heatmap showing associations between plasma p-tau levels and vascular risk factors at baseline on cognitive decline over time. Estimates reported correspond to the interaction between time and the marker of interest. All p-tau measures were included as continuous values **D.** For visualization purposes, participants were grouped into tertiles to show the effect of baseline p-tau levels on MoCA performance over time.

*AD, Alzheimer’s disease; CH, cognitively healthy; MCI, mild cognitive impairment; MoCA, Montreal Cognitive Assessment;* p-tau, phosphorylated tau; *RAVLT, Rey Auditory Verbal Learning Test; SCD, subjective cognitive decline; t, tertile (1, 2 or 3).* Statistical significance is indicated as follows: * *p* < .05; ** *p* < .01; *** *p* < .001.
